# Mapping T cell infiltration patterns in glioma tumor tissue

**DOI:** 10.1101/2025.06.25.25330286

**Authors:** Tiffaney Hsia, Ana K. Escobedo, Syeda Maheen Batool, Emil Ekanayake, Gavin P. Dunn, Bryan D. Choi, Bob S. Carter, Leonora Balaj

## Abstract

**Background:** The glioma immune repertoire has emerged as a vital point of interest, particularly in the context of immunotherapeutics development and as a key player for prognostic and diagnostic biomarker identification.

**Methods:** Tumor tissue was collected from glioma patients and targeted immune repertoire sequencing of tumor infiltrating lymphocytes (TIL) from each of the four collected glioma tumor subtypes was performed. Gliomas were stratified based on WHO21 classification to map the TCR landscape of astrocytomas (grade II/III, grade IV), glioblastomas, and oligodendrogliomas.

**Results:** Following stratification of TCR repertoires and complete clonotype, V-J cassette, and CDR3 analysis, we identified cohort-specific levels of diversity, clonotype sharing, and conservation. Partitioning of these repertoires based on TCR diversity revealed significant influence on patient survival. Furthermore, mapping of CDR3 binding regions to antigens and their origins highlighted prognostic biomarkers and identified sequences binding to viral signatures associated with patient clinical outcomes.

**Conclusion:** These findings underscore the importance of characterizing TCR repertoires in the context of the patient clinical condition. These unique repertoire signatures and correlated antigens may facilitate patient outcome prognostication and serve as a potential foundation for immunotherapeutic applications.

**Key Points:**

- Glioma subtypes can be differentiated via TCR repertoire characterization.
- T cell diversity in gliomas influences patient overall survival.
- TCR Clonotypes are binding sites for prognostic tumor proteins and viral disease.

**Importance of the Study:** In this study, we characterized the infiltrating immune repertoire of glial tumors through targeted T cell receptor sequencing of four glioma subtypes: grade II/III astrocytomas, grade IV astrocytomas, glioblastoma, and oligodendrogliomas. We performed multi-level analyses to examine TCR repertoire diversity across these subtypes, identifying unique clonotype prevalence and region usage in alpha-beta T cells. Classification of clonotypes relative to known associated antigen databases revealed unique signatures related to the viral diseases of cytomegalovirus and Epstein-Barr virus, as well as clonotypes with binding affinity for human tumor-derived proteins prognostic for glioma. This study reveals a unique perspective into the differentiation of glioma subtypes based on TCR clonotypes and further solidifies the role of TCR mapping in the patient care paradigm.

## INTRODUCTION

Gliomas comprise over 26% of all tumors in the central nervous system (CNS)^1^. They originate from glial cells and are highly heterogeneous, with diverse histopathologies and genetic compositions. Clinical outcomes have been shown to be dependent on the tumor molecular landscape and remain relatively poor. As such, it is critical to elucidate the tumor landscape and its interaction with the surrounding microenvironment to improve understanding of tumor behavior and its potential response to therapeutics. The role of the immune system in the tumor microenvironment (TME) has been of great interest, particularly in understanding T cell infiltration as it relates to tumor progression. Understanding this crosstalk elucidates key factors and targets that may be leveraged to develop immunotherapies, such as monoclonal antibodies^2^, vaccines, and adoptive T cell therapies^3^, to subsequently improve patient outcomes. Immunotherapies highly rely on infiltrative T cell function and structure. Therapeutic efficacy is therefore critically dependent on understanding and mapping the tumor infiltrating lymphocytes (TIL) of different glial tumor categories. While tumor evolution is highly complex and is influenced by an extensive network of variables, the clonotypic diversity of the T cell receptor (TCR) landscape plays an important role in anti-tumor immune response. Only a handful of studies have characterized the infiltrative TCR landscape in gliomas, with findings showing stratification of malignant brain tumors according to clinical grade^4^, mapping and identification of enriched TILs in glioma^5^. Additionally, characterization of the tumor and infiltrating T cell repertoire has revealed unique relationships between the tumor genomic landscape and immunologic diversity in malignant brain tumors^6,7^. Despite these advances, much further work is needed to understand the relationship between tumor histopathology and TCR repertoires to understand the role of endogenous immune response in the TME.

The immunological role of the T cell in the local TME relies heavily on TCR structure^8^. Specifically, the somatic recombination of the variable (V) and joining (J) regions and sandwiching of an encoding complementarity-determining 3 (CDR3) domain on the α chain and β chain loci facilitate specific target epitope binding. The unique structural configuration and binding affinity to different targets is driven by CDR3 sequence diversity. Adaptive immune response in the TME is thereby directly dependent on T cell infiltration, activation, and clonal expansion. As such, mapping of the infiltrative TCR repertoire of glial tumors is essential to providing insight into TME immunological activity and enables functionalized characterization with respect to the patient clinical condition.

In this study, we sought to characterize the TCR repertoires of infiltrating T cells in glial tumors across different disease subtypes. Using tumor tissue samples, we performed targeted TCR sequencing to characterize infiltrating TCR clonotypes across cohorts. Furthermore, clinico-pathological conditions were integrated in the analysis which revealed correlations between clinical variables and the immune repertoire of the glioma microenvironment.

## METHODS

### 1. Ethics approval and consent to participate

This study was conducted in compliance with the Declaration of Helsinki and with the International Conference on Harmonization Good Clinical Practice Guidelines protocol. Written and informed consent was obtained from patients undergoing surgery at Massachusetts General Hospital (MGH) after briefing on the nature of the study and the potential associated risks. The study was approved by the Internal Review Board ethical committee at MGH (2017P001581).

### 2. Tumor tissue processing

Tumor tissue was collected and transferred to a sterile vial immediately following surgical resection. Samples were dissected on a sterile 100 mm plate with a sterile scalpel into equal mass aliquots, flash frozen, and stored at -80°C.

### 3. RNA extraction from tumor tissue

Tumor tissue was retrieved from storage at -80°C and placed on a 100 mm sterile plate for weighing. Tissue was then completely dissociated using a 32 gauge needle in dissociation buffer and RNA was immediately extracted using the RNeasy Kit (Qiagen; Hilden, Germany) according to manufacturer’s recommendations.

### 4. Library preparation

Complementary DNA (cDNA) libraries were prepared using the QIAseq Immune Repertoire RNA Library Prep Kit Panel (Qiagen; Hilden, Germany) according to the recommended protocols. Quality control (QC) was performed to confirm initial RNA quality and quantity and final library quality and quantity. Pre-preparation input RNA was characterized using the BioAnalyzer High Sensitivity RNA Pico Assay (Agilent Technologies; Santa Clara, CA). cDNA libraries were quantified using the BioAnalyzer High Sensitivity DNA Assay (Agilent Technologies; Santa Clara, CA) and the QIAseq Library Quant Assay Kit (Qiagen; Hilden, Germany) in conjunction with a StepOne Real-Time PCR system (ThermoFisher Scientific; Waltham, MA). PCR conditions followed those described in the QIAseq Immune Repertoire protocol. All BioAnalyzer assays were quantified using the 2100 BioAnalyzer Instrument (Agilent Technologies, Santa Clara, CA). Libraries were pooled at equal molar concentration and a final quantification was performed via PCR prior to sequencing.

### 5. MiSeq Sequencing

Pooled libraries were denatured using fresh 0.2 N NaOH for 5 minutes and quenched with ice cold Hybridization Buffer from the MiSeq Reagent Kit v3 (Illumina; San Diego, CA). The sequencing cartridge (600 cycle) was prepared and the pooled, denatured library was loaded according to manufacturer’s instructions for sequencing.

### 6. Data analysis

Initial QC of the sequencing runs were viewed and confirmed in Basespace. Subsequently, all FASTQ derived from sequencing were demultiplexed and analyzed using Qiagen CLC Workbench using the Immune Repertoire pipeline, in accordance with the default values. TCR characteristics were characterized using CLC workbench. Diversity of the CDR3 sequences were calculated in R using several packages including vegan^9^ and ImmuneArch^10^, package use as detailed in figure legends. The ImmuneArch package was used for downstream analyses of clonotypes as well. Clustering analysis of productive sequences was performed using the GLIPH2^11^ web user interface (http://50.255.35.37:8080/) with the “version 2” clustering algorithm, and clusters were evaluated for significance according to the following parameters: number of subjects ≥3, number of unique CDR3s ≥3, vb score ≤0.05, and length score ≤0.05. Clusters were subsequently ordered for significance according to the final_score and plotted using the R package igraph^12^. Sankey plots were plotted using SankeyMATIC^13^ and motif conservation logos were drawn using the WebLogo online user interface^14^. All *k*-mer motif analysis was performed using ImmuneArch and peptide properties were identified using PepDraw (https://www2.tulane.edu/∼biochem/WW/PepDraw).

### 7. Statistical analysis

Statistical testing was performed using unpaired student’s t-tests as well as one-way and two-way ANOVAs, unless otherwise stated, with significance defined at a *p*-value of less than or equal to 0.05. Heteroscedasticity of populations was considered for all t-tests and analysis was performed accordingly.

## RESULTS

### TCR repertoires from glial tumors reveal disease-specific trends in expression

In an effort to characterize the T cell repertoire of various glial tumor disease states, we performed bulk, targeted T cell receptor (TCR) sequencing of tumors from patients with IDH1 mutant (IDH1mut) grade II/III astrocytomas (*n* = 9), IDH1mut grade IV astrocytomas (*n* = 3), IDH1 wildtype (IDH1wt) glioblastoma (GBM, *n* = 7), and oligodendrogliomas (*n* = 3). Cohorts were designed such that patient ages spanned similar ranges and each cohort was categorized according to WHO21 classification. To assess the robustness of this bulk sequencing method, clonotypes from matched chains were compared, which showed near equal characterization of alpha and beta chain-derived clonotypes (alpha:beta, gamma:delta; **Fig. 1A**). All detected clonotypes were subsequently stratified based on clonotype productivity as defined by the CDR3 sequences into clonotypes that were (1) productive, (2) out of frame, or (3) had a premature stop codon (**Fig. 1B**). Across all patients, we identified that the majority of clonotypes had productive sequences for epitope binding regardless of disease. Henceforth, analysis was performed on productive clonotypes alone. Unique clonotypes, as defined by a unique combination of variable region, joining region, and CDR3 sequence, were then compiled to produce a summary of all productive, unique clonotypes across all four chains (TRA, TRB, TRG, and TRD). This comparison revealed unique patterns of clonotype prevalence in glioma tumor subtypes. Grade IV astrocytomas demonstrated a broader range and a higher average of unique clonotype counts as compared to grade II/III astrocytomas, glioblastomas, and oligodendrogliomas (**Fig. 1C, D**). All productive, unique clonotypes were subsequently ranked in order of prevalence (“count”), as identified via sequencing, and clonotypes from alpha (TRA) and beta (TRB) chains (**Fig. 1E**) and delta (TRD) and gamma (TRG) chains (**Supp. Fig. 1A**) to elucidate the prevalence of highly expanded clones, mid-range clones, and singletons (clonotypes with a single read). The number of singleton clonotypes, as defined by range and mean, were assessed per cohort and chain (**Fig. 1F**, **Supp. Fig. 1B**). The grade IV astrocytoma cohort yielded a higher number of singletons across all chains, reflective of the diversity of productive, unique clonotypes seen in **Fig. 1C**. A breakdown of the complete clonotype into variable and joining regions, in fact, revealed cohort-unique clustering of the variable (**Fig. 1G**) and joining (**Fig. 1H**) region usage across alpha, beta chains. Independent plotting of variable and joining regions, known as a V-J cassette, across cohorts demonstrated regions of cohort-dependent expression (**Fig. 1I**) with the highest average expression, prevalence, and range of use in both the alpha and beta chain cassettes found in the grade IV astrocytoma cohort. Analysis of the top 10 most commonly occurring V-J cassettes from TRA and TRB chains in each cohort revealed overlapping usage of unique variable and joining regions within each subtype and across cohorts (**Fig. 1J**). In fact, the two astrocytoma cohorts were found to have overlap in TRA variable region 4 and TRB regions V-19 and V-20-1 within their most expressed V-J cassettes. Interestingly, other instances of usage overlap were seen between grade II/III astrocytomas, glioblastomas, and oligodendrogliomas, exclusive of grade IV astrocytomas, with TRA V-19, and V-13-1. This overlap in the top ten V-J cassette usages across cohorts demonstrates TCR structural similarities across glial tumor subtypes.

**Figure 1.**
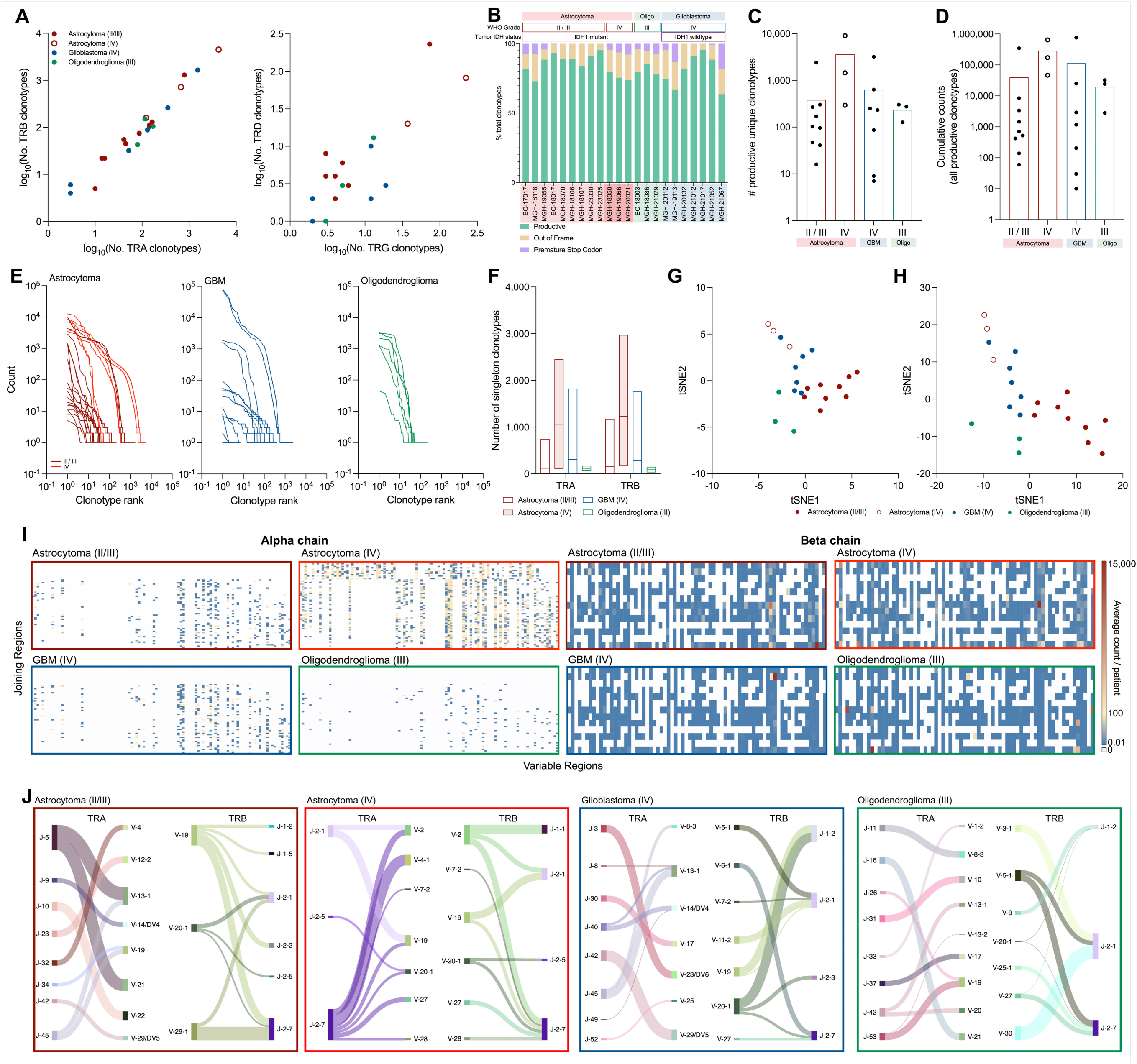
Inter-tumoral classification of TCR clonality. RNA from bulk tumor tissue from patients with astrocytoma grade II/III (*n* = 9), astrocytoma grade IV (*n* = 3), glioblastoma grade IV (*n* = 7), and oligodendrogliomas grade III (*n* = 3) was extracted for targeted TCR sequencing. Log-normalized quantification of alpha chain (TRA) and beta chain (TRB) derived total clonotypes was compared to assess sampling rigor (**A**, TRA:TRB r^2^ = 0.9553, TRG:TRD r^2^ = 0.6891). Detected clonotypes were stratified based on productivity (productive, out of frame, and premature stop codon) based on CDR3 amino acid sequences from TRA, TRB, TRG, and TRD chains (**B**). Productive, unique clonotypes were identified from TRA, TRB, TRG, and TRD chains from all patients (**C**). Total number of clonotype counts were determined for each patient according to disease category (**D**). All productive clonotypes were given a rank position as defined by order of descending frequency in each sample in each TRA and TRB condition. Clonotype rank was plotted with respect to frequency (**E**). Singletons, as classified by a count frequency of 1, were quantified per patient, per disease (**F**). T-distributed stochastic neighbor embedding was performed on all detected, productive variable regions (TRA, B, G, D) to assess similarity of unique variable region detection within disease categories, across all patients. Following multiple iterations, final tunable parameters were defined: perplexity of 5, max iteration of 1500 (**G**). Joining regions from productive clonotypes of all four chains were additionally analyzed via t-SNE (**H**). Average count per patient of all productive V-J cassettes across disease were analyzed for abundance across disease (**I**). The top 10 most commonly occurring V-J cassettes across all patients in each cohort were identified and the relationship between variable and joining regions were determined with respect to expression level (as defined by flow thickness) within each disease condition (L to R: grade II/III astrocytoma (dark red box), grade IV astrocytoma (light red box), GBM (blue box), oligodendroglioma (green box); **J**). Sankey plots were plotted using SankeyMATIC (sankeymatic.com).

### Tumor type affects both the diversity and similarity of T cell receptor (TCR) profiles

To investigate the degree of similarity between patient TCR repertoires (V, J, CDR3), we quantified population overlap using Morisita’s overlap index, in which larger values represent increased levels of similarity (**Fig. 2A**). We observed patterns of similarity within patient cohorts, patient-unique signatures, and regions of cross-subtype similarities as evidenced by grade II/III astrocytomas, grade IV astrocytomas, and GBMs (**Fig. 2A**). A further breakdown of clonotypes to evaluate CDR3 amino acid sequences alone revealed cohort-based stratification by average Renyi diversity, with the grade IV astrocytoma cohort exemplifying a higher average diversity over glioblastomas, grade II/III astrocytomas, and oligodendrogliomas (**Fig. 2B**). The point at which Renyi diversity collapsed for all four cohorts (Hill = 1, Shannon Diversity) was chosen for additional characterization specific to TRA-derived and TRB-derived CDR3 sequences (**Fig. 2C**). While the range of Shannon diversity across all patients was similar, the average diversity and diversity distribution per patient cohort was found to be higher in grade IV astrocytomas patients relative to the other three cohorts. We then assessed CDR3 amino acid length for alpha and beta chains to determine the range and prevalence of lengths (**Fig. 2D**). As expected, average and median amino acid lengths of all chains were found to be similar. However, evidence of significantly longer CDR3 clonotypes were found in patients with grade IV astrocytomas, suggesting variations in epitope binding behavior. Comparison of unique CDR3 sequences from each cohort elucidated sequences common and unique to each category (**Fig. 3E**). The average expression per patient of the 59 common sequences between grade IV astrocytomas (IDH1mut) and GBM (IDH1wt) was assessed and revealed strikingly opposite patterns of expression between the two subtypes (**Fig. 3F**). Of the 53 common sequences (**Fig. 3G**), four CDR3 clonotypes demonstrated nearly equal levels of average expression across the three cohorts. Further analysis of the top 10 most expressed CDR3 sequences (average count per patient) further attested to the sharing of GBM TCR clonotypes in the two other cohorts (**Fig. 3H**). Within the top 10 most expressed sequences identified in the GBM cohort, these sequences were also identified in the grade II/III astrocytoma, grade IV astrocytoma, and oligodendroglioma cohorts, albeit at lower average expression levels. It is interesting to note that the top 10 most expressed sequences found in the other three cohorts (grade II/III astrocytoma, grade IV astrocytoma, and oligodendroglioma cohorts) were not as frequently found across disease subtypes.

**Figure 2.**
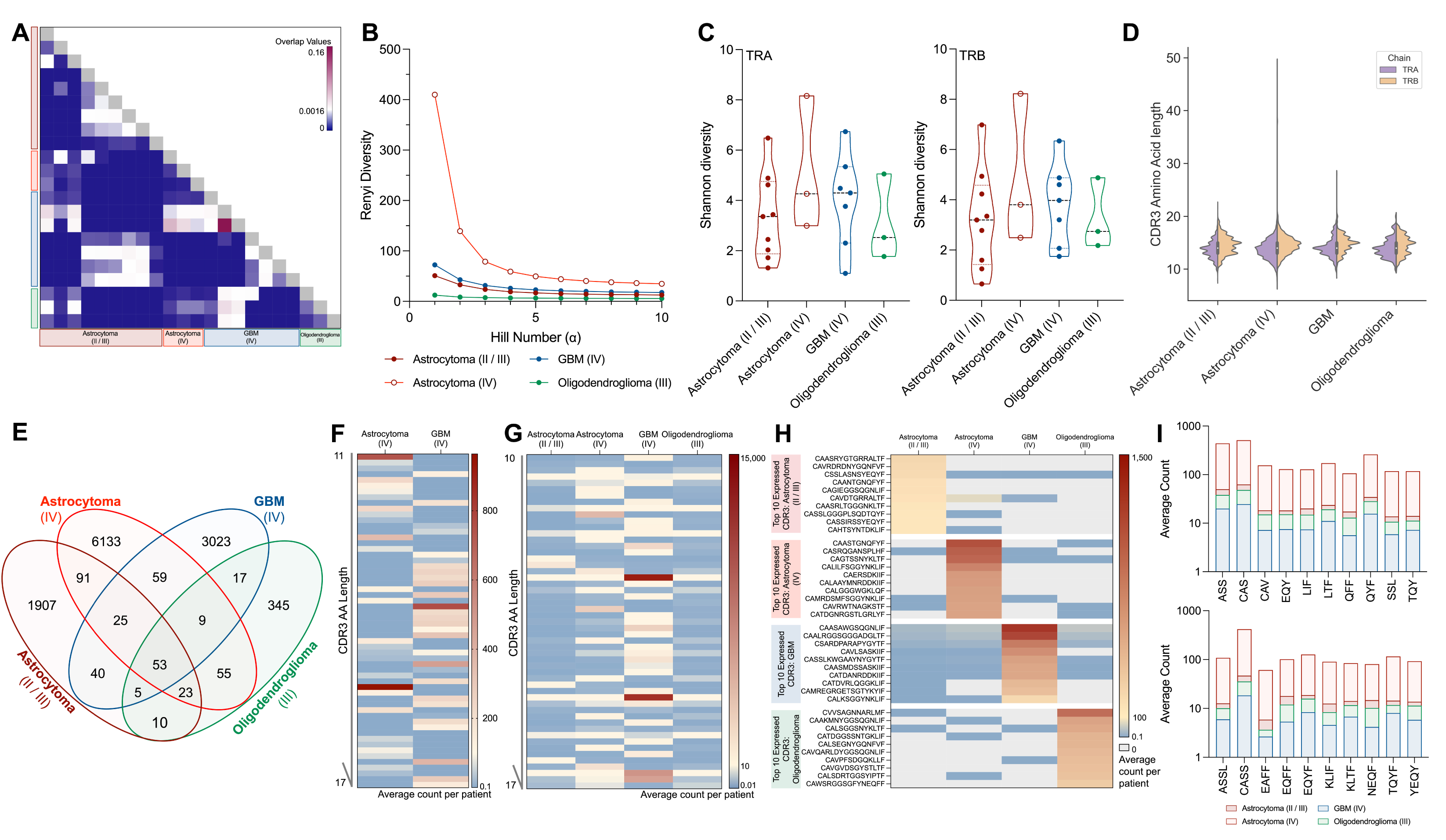
Evaluation of productive TCR diversity. A comprehensive (V chain, J chain, CDR3 amino acid sequence) assessment of sample similarity within disease was first determined using Morisita’s overlap index (**A**). CDR3 amino acid sequence diversity was analyzed via Renyi diversity profiling (**B**). A patient-by-patient Shannon diversity based on CDR3 sequences from the alpha chain and beta chain was calculated (**C**). Amino acid sequences from the alpha and beta chains from each patient were compiled to demonstrate distribution of CDR3 lengths (inclusive of CASS motif) across chains and across disease (**D**). Productive CDR3 sequences from alpha and beta chains were compared across disease to identify overlap and unique expression (**E**). The average expression per patient of the 59 common genes between grade IV astrocytoma and GBM were compared (**F**) as well as the expression of the 53 genes common across all 4 cohorts (**G**). The top ten most expressed CDR3 sequences in each disease group (across all patients within the disease category) were identified and their average expression per patient was calculated for each category (**H**). Motif frequency analysis of CDR3 sequences across cohorts via the top 10 most abundant 3-mer and 4-mer sequences (**I**).

**Figure 3.**
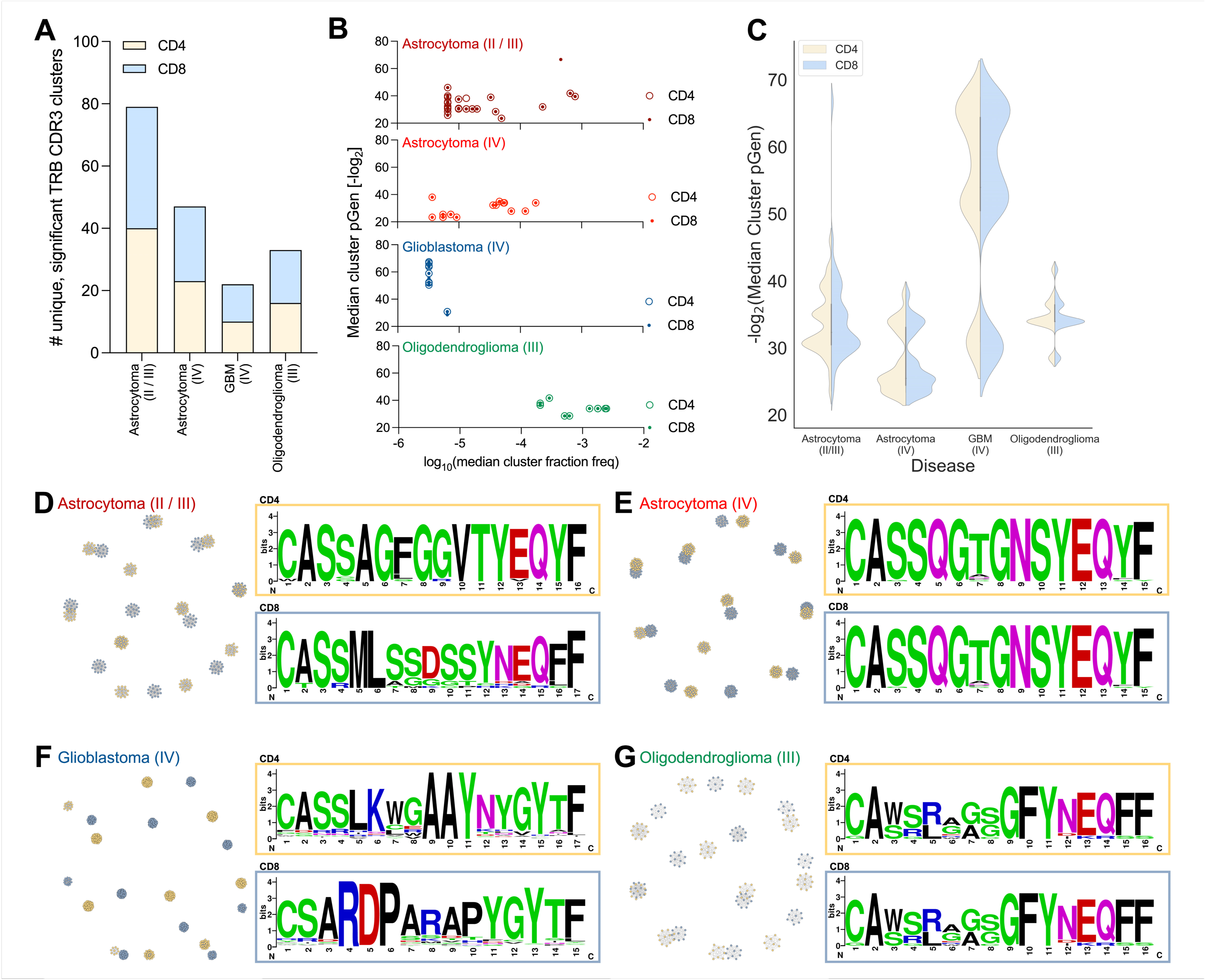
CDR3 sequence mapping reveals patterns associated with disease. Peptide antigen prediction from detected TRB CDR3 sequences was performed via GLIPH2 analysis. Number of unique and statistically significant T cell clusters was obtained relative to CD4 and CD8 TCRs with respect to disease category (**A**). Median frequency (normalized to total counts) and generation probability (pGen) scores for each unique, significant cluster was calculated for each disease and T cell dataset (**B**). Comparative view of pGen scores for all unique, significant clusters associated with CD4 T cells as compared to CD8 T cells for each disease (**C**). Network clustering of the top 10 largest unique, significant clusters from CD4 and CD8 CDR3 sequence enrichment analysis was conducted for patients with grade II/III astrocytomas (**D**), grade IV astrocytomas (**E**), GBM (**F**), and oligodendrogliomas (**G**). Amino acid motif conservation of the largest significant cluster from CD4 (yellow) and CD8 (blue) datasets was performed using WebLogo.

We further sought to explicate the convergence of the productive CDR3 sequences at the motif level through overlapping k-mer decomposition. This approach, which allows for identification of patterns of conservation within and across cohorts, facilitates functionalization of sequence motifs for epitope binding affinity prediction. We developed 2-mer, 3-mer, 4-mer, 5-mer, and 10-mer frequency matrices and isolated the top 10 most abundant sequences across all patients to examine cohort-enriched motifs (**Fig. 3I**, **Supp. Fig. 2**). We observed adherence to standard conservation of the CAS/CASS motif, commonly found at the beginning of CDR3 sequences. We also identified 3- and 4-mers which demonstrated relative enrichment in the grade IV astrocytoma cohort over other glioma subtypes. Amino acid characterization of these most abundant 4-mer motif for grade IV astrocytomas (EAFF) revealed retention of aromatic molecular structures (namely phenylalanine), residues important to structural stability and protein-protein interactions^15^.

### Conservation and clustering of CDR3 sequences distinguish tumor cohorts

We next performed TCR annotation and clustering analysis and calculated the generational probability of remarkable clonotypes with respect to naive CD4 and CD8 repertoires (TRB database). Given that bulk TIL sequencing was performed, we have referenced our TCR repertoires to both CD4 and CD8 references to comprehensively map the CDRR3 motifs. To predict antigen specificity of CDR3 sequences relative to known CD4 antigens and CD8 antigens and identify conserved CDR3 sequences within each cohort, we used the Grouping of Lymphocyte Interactions by Paratope Hotspots v2 (GLIPH2) algorithm to cluster sequences based on their specificity and motif similarity within each cohort-based dataset (**Fig. 3A**)^11^. The number of unique, significant clusters as matched to both CD4 and CD8 datasets were assessed for cluster density as well as generational probability. Using the Optimized Likelihood estimate of immunoGlobulin Amino acid sequences (OLGA) algorithm, the generational probability (pGen) of all unique significant clusters were calculated and Log2 transformed for plotting purposes (**Fig. 3B,C**)^16^. The calculated pGen of each cluster provides insight into the publicity of the clonotypes, with low pGen clusters representing clonotypes not widely shared across the cohort and high pGen clusters indicative of convergence and high levels of sharing^17^. This analysis showed that the GBM cohort had the densest population of clusters and that the grade IV astrocytoma cohort had the highest pGen (low -log_2_(pGen)) out of all the cohorts. Clonotype sharing (pGen) explored by way of violin plots revealed a relatively increased skewing of clonotype generational probability towards a low pGen within the GBM cohort. Through network clustering of the top 10 largest clusters (CD4: yellow, CD8: blue), we found significantly denser clusters in the GBM cohort as compared to other categories (grade II/III astrocytoma: **Fig. 3D**, grade IV astrocytoma: **Fig. 3E**, GBM: **Fig. 3F**, oligodendrogliomas: **Fig. 3G**). Clustering was performed using the Fruchterman-Reingold layout algorithm. Breaking down each cluster (10 largest) to examine cluster sharing across patients, we identified levels of sharing with respect to the number of patients expressing the clonotype unique to each cluster (**Supplementary Table 1**).

In order to further examine cluster characteristics, we explored the conservation of the CDR3 sequences within the largest cluster. Motif mapping of the largest clusters revealed close resemblance of conservation across the naive CD4 and CD8 databases, with unique specificity groups in the astrocytoma and other glial tumor cohorts. Characterization of the conserved amino acids of each CDR3 sequence further demonstrates cohort-unique structural elements that may influence epitope binding.

### Using TCR repertoires as prognostic indicators through clinico-pathological correlation and antigen mapping

We next sought to assess the relevance of the TCR repertoires relative to the patient clinical condition. To first assess known confounding factors, we first examined tumor characteristics such as tumor methylation or mutation status and patient characteristics relative to patient survival (**Supplementary** Fig. 3A-D). When we binned overall survival into three categories (0-12, 12-60, and 60+ months) and quantified the average number of unique clonotypes per patient in each tumor subtype, we found an upward trend in clonotype prevalence within the grade II/III astrocytoma and GBM cohorts (**Supp. Fig. 3E**), while oligodendrogliomas (*n* = 2 patients per bar) showed a relatively equal distribution. We then calculated the Shannon Diversity by combining TRA and TRB profiles of each patient and correlated it with tumor size (grade II/III astrocytoma: red, grade IV astrocytoma: red open circle, GBM cohort: blue, oligodendroglioma: green). Irrespective of subtype, we observed a negative trend of repertoire diversity with respect to the tumor volume (**Fig. 4A**), as determined by imaging. Based on Shannon Diversity, we stratified all patients into categories of high and low diversity (median-split, high diversity >4.15 and low diversity ≤4.15). Notably, a substantial difference was observed between the overall survival probabilities of patients with high TCR repertoire diversity and low TCR repertoire diversity (*p* = 0.074; **Fig. 4E**). Comparison of progression free survival (PFS) between high and low diversity groups revealed a significant difference in the survival probability (*p* = 0.0491; **Fig. 4F**), with patients with low TCR diversity showing an improved PFS of 145.7 months as compared to 10.17 months (median PFS). To consider additional confounding factors of survival, the number of patients with tumor MGMT methylation, a prognostic marker for glioma OS, was determined. Incidentally, the number of patients with unmethylated and methylated MGMT in each of the low diversity (6 to 4, respectively; N/A = 1) and high diversity (5 to 4, respectively; N/A = 2) groups were relatively similar, such that methylation status may not influence diversity-stratified patient survival.

**Figure 4.**
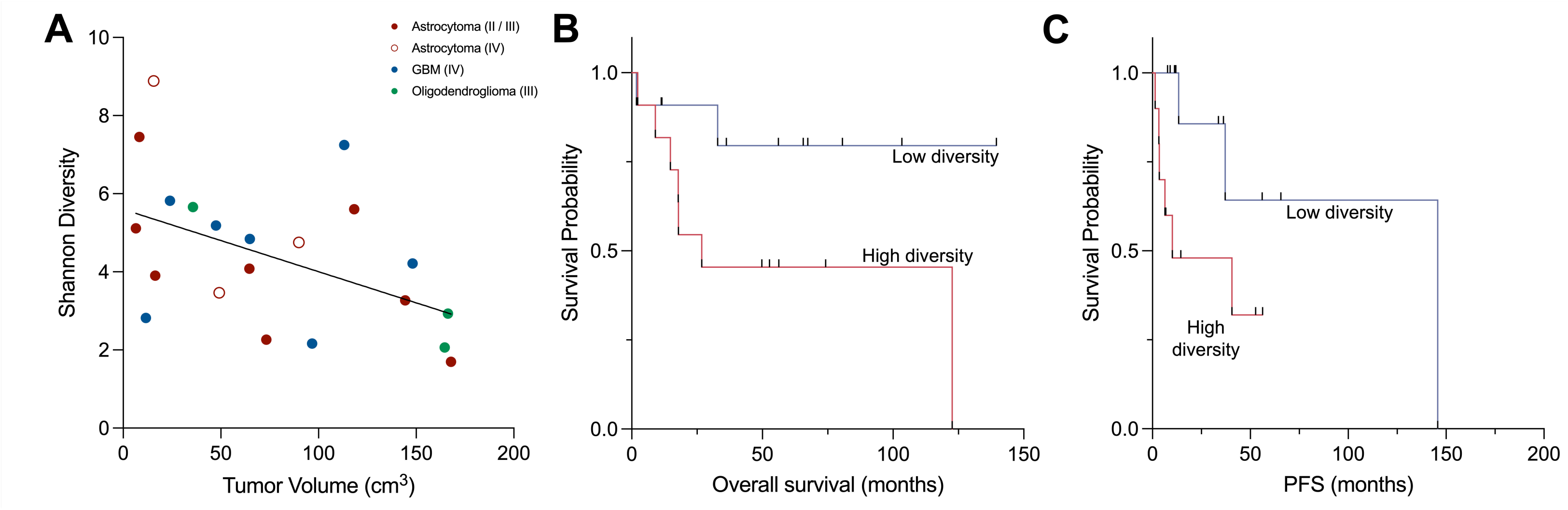
Clinicopathological comparison of cohorts relative to alpha-beta chain clonotypes. Clinical outcomes and tumor pathology was assessed relative to the cohorts and the detected clonotypes. Tumor volume (cm^3^) was compared to calculated Shannon Diversity of the combined TRA+TRB clonotypes (**A**; grade II/III astrocytoma: red, grade IV astrocytoma: open red circle, GBM: blue, oligodendroglioma: green). All TCR repertoires were median-split into high and low diversity (cut-off = 4.15, inclusive in the low diversity population). Overall survival curves for all patients with high (red) and low (blue) CDR3 diversity (**B**; log-rank p = 0.074). Progression free survival curves for all patients with high (red) and low (blue) CDR3 diversity (**C**; log-rank *p* = 0.0491).

We then mapped TRA and TRB profiles to the human VDJ database (VDJdb)^18^, which enables identification of antigens from previously reported CDR3 clonotypes. Of the CDR3 clonotypes detected in our patient population, we identified a fraction of clonotypes with previously reported antigen specificity (grade II/III astrocytoma: 5.9%, grade IV astrocytoma: 4.8%, GBM: 7.6%, oligodendroglioma: 3.3%; **Fig. 5A**). From these annotated clonotypes, we identified antigens of various origins including cytomegalovirus (CMV), Epstein-Barr virus (EBV), human, and influenza A. Interestingly, influenza A derived antigens were found to overlap with antigens targeted in vaccinations^19^. Within antigens of human origin, we found several had multiple unique CDR3 sequences linked to the same antigen (**Fig. 5B**; grade II/III astrocytoma - dark red, grade IV astrocytoma - light red, GBM - blue, oligodendrogliomas - green). These antigens, bone marrow stromal cell antigen 2 (BST-2; *p* = 0.031) and melan-A (MLANA; *p* = 0.035), were also found to be prognostic for glioma OS based on low and high expression profiles (based on “best expression cutoff” as determined in the Human Protein Atlas; **Fig. 5C**)^20^. We further identified annotated antigens of viral origin. The two most common pathologies, cytomegalovirus (CMV) and Epstein-Barr Virus (EBV), were found in a large proportion of patients, with clonotypes for CMV found in 21 out of 23 patients sequenced (**Fig. 5D**). We next assessed the relevance of viral co-expression within each patient cohort and found that clonotypes for both CMV and EBV were expressed in all three tumor subtypes, ranging from 44% to 75% (**Fig. 5E**). Interestingly, the two most enriched epitopes (immediate early protein I - IE1, and pp65) were found to have significantly higher counts of antigen-specific clonotypes in grade IV astrocytomas followed by GBM, grade II/III astrocytomas, and oligodendrogliomas (**Fig. 5F**). Distribution of the total number of clonotypes revealed an overwhelming prevalence (∼50% for all astrocytomas, 37% for GBMs, and 43% for oligodendrogliomas) of mapped CDR3 sequences binding to CMV-derived antigens **(Fig. 5G**). Similarly, epitopes derived from EBV were found in patients across all three cohorts. Antigens with high levels of expression (EBV immediate early protein - BZLF1, EBV nuclear antigen 3A - EBNA3A, and EBV nuclear antigen 4 - EBNA4) also reported higher counts of clonotypes in GBM followed by astrocytomas and other glial tumors (**Fig. 5H**).

**Figure 5.**
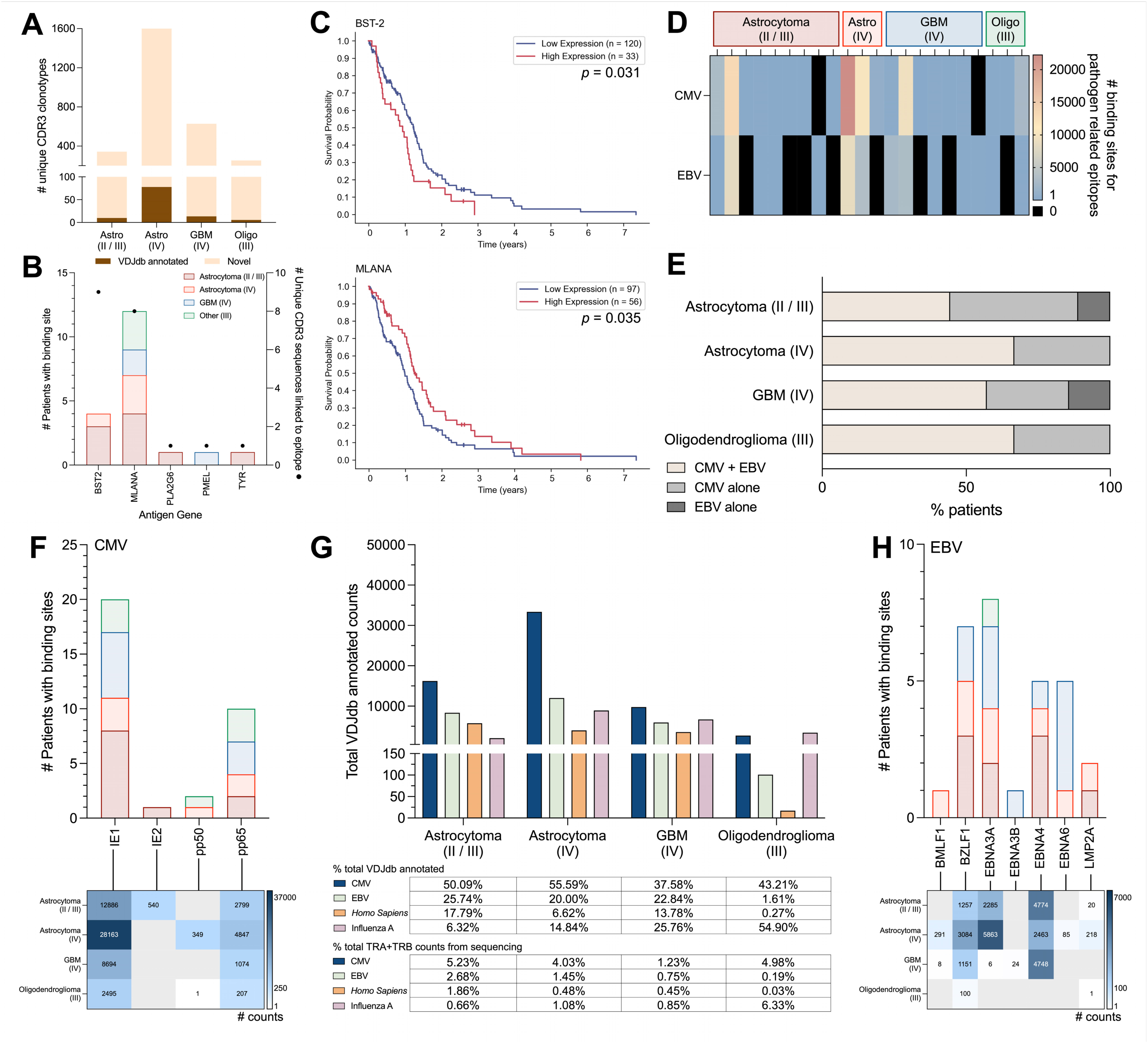
Pathological profiling of CDR3 sequences relate to clinical outcomes. TRA and TRB derived CDR3 sequences were annotated using the VDJ Homo Sapiens database (VDJdb) to identify antigen genes (**A**). All human-derived antigen genes (**B**) were examined in the context of glioma for prognostic significance (**C**). Survival analysis, expression cut-offs (best expression cutoff), and subsequently calculated p-values were identified according to the Human Protein Atlas database. CDR3 sequences were also assessed against CMV antigen gene targets and the number of patients bearing receptors as well as total number of binding sites found across all patients was quantified (**D**) and patients with clonotypes for multiple pathologies were identified (**E**). Binding site frequency for unique Cytomegalovirus (CMV) epitopes (**F**) were assessed per disease subtype. All clonotypes annotated using VDJdb were quantified as defined by the number of counts per clonotype. Total counts of all VDJdb annotated counts from all patients in each cohort were summed and plotted for CMV, EBV, Homo Sapiens, and Influenza A (**G**). Percentages of clonotypes annotated with VDJdb relative to total VDJdb annotated and total sequenced TRB clonotypes were calculated as well. Binding site frequency was also determined for unique Epstein-Barr Virus (EBV) epitopes (**H**).

## DISCUSSION

Here, we use targeted TCR repertoire sequencing to map the immunological landscape of glioma tumor subtypes (grade II/III astrocytoma, grade IV astrocytoma, GBM, and oligodendroglioma) in concordance with WHO21 classification criteria. We identify a new trend in the evolution of IDH1mut astrocytoma TCR repertoire diversity, and distinguish grade IV astrocytomas from grade IV glioblastoma, grade II/III astrocytoma, and oligodendroglioma via TCR clonotypes. Through a global analysis of complete clonotypes and more comprehensive analysis of V-J cassettes and CDR3 motifs, we identified tumor-related signatures of immune response. Our findings also revealed clinical and pathological correlations with the TCR repertoires, suggesting T cell infiltration plays a critical role in overall patient outcomes. Previous studies on TCR repertoires have reported grade-based stratification of immunological phenotypes, with glioblastoma and metastatic tumors demonstrating increased numbers of unique clonotypes and increased diversity as compared to low grade gliomas^4,21–23^. Here, we performed multi-level comparative analysis of TCR repertoires from grade II/III astrocytomas, grade IV astrocytomas, GBMs, and oligodendrogliomas. We found increased unique clonotype abundance and higher clonotype diversity in grade IV astrocytoma (IDH1mut) tumor tissue compared to other disease categories, even GBM (IDH1wt). Interestingly, we discovered elevated levels of singleton clonotypes across all grade IV astrocytoma patients as well, suggesting increased richness of the TCR repertoire. When comparing the similarity of CDR3 amino acid sequences across the four cohorts, the high diversity of the grade IV astrocytoma cohort enabled overlap with the other two cohorts. In particular, we identified higher levels of overlap between the grade II/III and grade IV astrocytoma cohorts, supporting an evolutionary trajectory of astrocytoma to higher WHO disease grade. Further study of TCR repertoires is required to understand the course of T cell activation, and clonotype expansion and conservation in disease evolution. In comparison, despite the relatively lower diversity found in GBM, conservation of the top ten most common GBM-derived CDR3 binding sequences was identified across all disease cohorts, suggesting the influence of additional TME and intrinsic tumor factors on the composition of TILs.

Recent studies have shed light on the role of sequence conservation in T cell epitope binding^24^. To expand on this paradigm, we further explored CDR3 amino acid sequences through *k*-mer conservation study. We assessed cohort similarity using sliding *k*-mer windows of lengths 3 through 10. We identified distinct sequences across cohorts as well as evidence of global conservation such as with the commonly found CASS motif at the beginning of CDR3 sequences^25^. *K*-mer motif analysis, particularly through amino acid characterization, functionalizes particular regions of the CDR3, providing increased understanding of binding affinity on the exposed CDR3 domain of the TCR. Through complete CDR3 analysis, we found high conservation of repeat motifs in grade IV astrocytomas, which may facilitate target identification. The majority of complete clonotypes (CDR3 sequences) identified within the GBM cohort were publicly shared. However, we also found a selection of private clonotypes, suggesting T cell response to a specific trigger unique to these patients.

TCR repertoires have been previously reported as prognostic markers for patient survival in response to immunotherapy in gliomas and other cancers^26–28^. Interestingly, analysis of TCR diversity of the cohort sequenced in this study revealed a link between Shannon diversity and survival probability (overall and progression free) across all patients regardless of disease subtype. Additionally, we identified antigens to CDR3 sequences from our sequencing study, BST-2 and MLANA, that are prognostic for overall survival. The correlation of the TCR repertoire, particularly CDR3 motifs, to known prognostic antigens paves the way for a deeper, more refined understanding of glioma evolution and T cell infiltration. These findings further suggest the importance of characterizing the TCR repertoire in patient clinical care. Further, we identified CDR3 sequences with epitopes of viral origin. The two most common pathologies, cytomegalovirus (CMV) and Epstein-Barr virus (EBV), have been long associated with glioma etiology and progression through both causative and associative means^29–31^. In fact, *in vitro* and clinical studies have capitalized on the presence of CMV antigens in the tumor to develop CMV-specific T cells for T cell therapy, finding increased localization of CMV-specific T cells at the site of antigen expression^32,33^ and evidence of immune activation following CMV protein vaccination^34^. Identification of abundant epitopes thus becomes advantageous to the development of new viral protein-specific T cells for immunological targeting.

Overall, the results of this study reaffirm the importance of elucidating the infiltrating TCR repertoire of gliomas. Through V-J cassette characterization and CDR3 amino sequence assessment, we report unique signatures of diversity related to glioma tumor subtype. Importantly, we have discovered evidence of CDR3 sequence conservation and clonotype expansion unique to each cohort, which highlight the unique epitope binding properties of these repertoires. These distinctions, in addition to clinico-pathological and viral signature correlation, enable identification of distinct biomarkers of tumor subtypes and provide prognostic insight relative to the immunological landscape. This investigation of the infiltrative T cell repertoire across glioma subtypes offers a new perspective into clinically-correlated TCR repertoires, contributes to the development of prognostic markers of disease progression based on immunological phenotypes, and supplements the growing field of T cell therapeutics against gliomas.

## Data Availability

All data produced in the present study are available upon reasonable request to the authors

## Acknowledgments

We would like to thank the patients and their families for their participation in this study.

## Data Availability Statement

Data and code used in manuscript preparation are available upon reasonable request.

## Ethics statement

Our study was conducted in accordance with principles for human experimentation as defined in the U.S. Common Rule and was approved by the Human Investigational Review Board of the Massachusetts General Hospital study center under IRB-approved protocol number 2017P001581. Samples were collected from patient population undergoing treatment at Massachusetts General Hospital.

## Data Availability Statement

All data produced in the present study are available upon reasonable request to the authors.

## Authorship

L.B., B.S.C., T.H. conceived the study; L.B. and T.H. designed the experiments; T.H. and A.K.E. performed the experiments; S.M.B. assisted with experiments; E.E. consented and enrolled volunteers to the study and collected and processed specimens; T.H. performed data analysis and interpretation; L.B. and B.S.C. reviewed the data and discussed findings; L.B., B.D.C., G.P.D., and B.S.C. reviewed and edited the manuscript; T.H. and L.B. wrote the paper. All authors revised and approved the manuscript for publication.

