## Supplemental Figures for "Mapping T cell infiltration patterns in glioma tumor tissue"

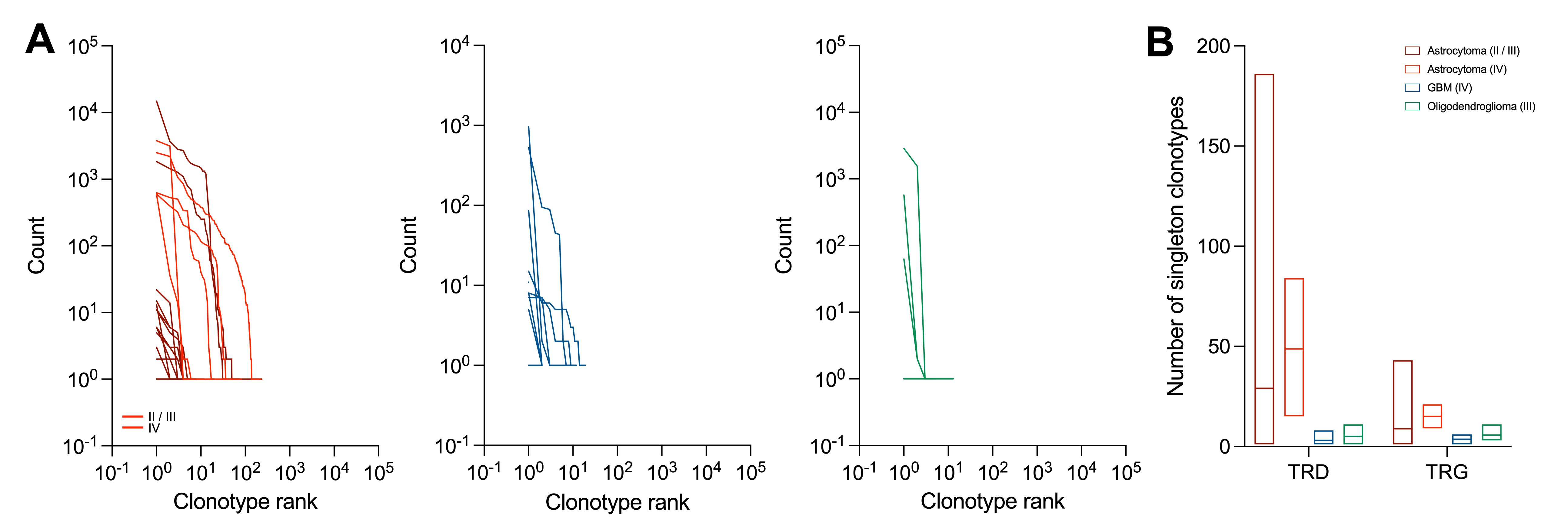

**Supplementary Figure 1. Ranking-based classification of TRG and TRD-derived productive clonotypes.** Clonotypes were ranked according to prevalence for patients in the astrocytoma (red), GBM (blue), and other glial tumor (green) cohorts (**A**). The number of singleton clonotypes were characterized per chain and cohort (**B**).

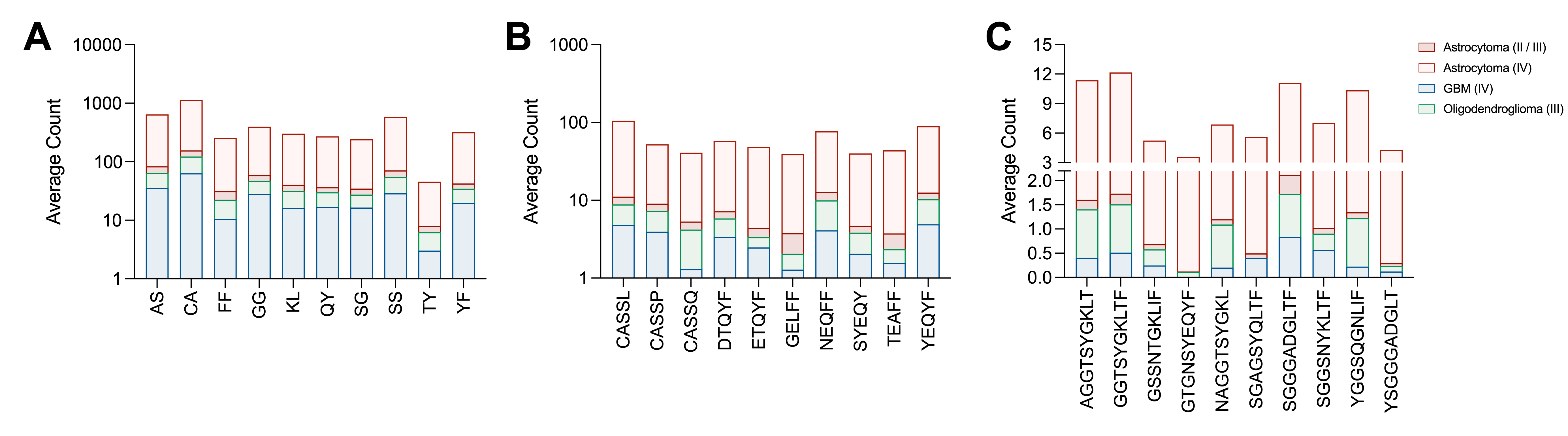

**Supplementary Figure 2.** **K-mer motif analysis of productive CDR3 sequences.** 2-mer (**A**), 5-mer (**B**), and 10-mer (**C**) representations of the CDR3 repertoires from each cohort were created. The top 10 most abundant motifs from each analysis were plotted and the average count reported.

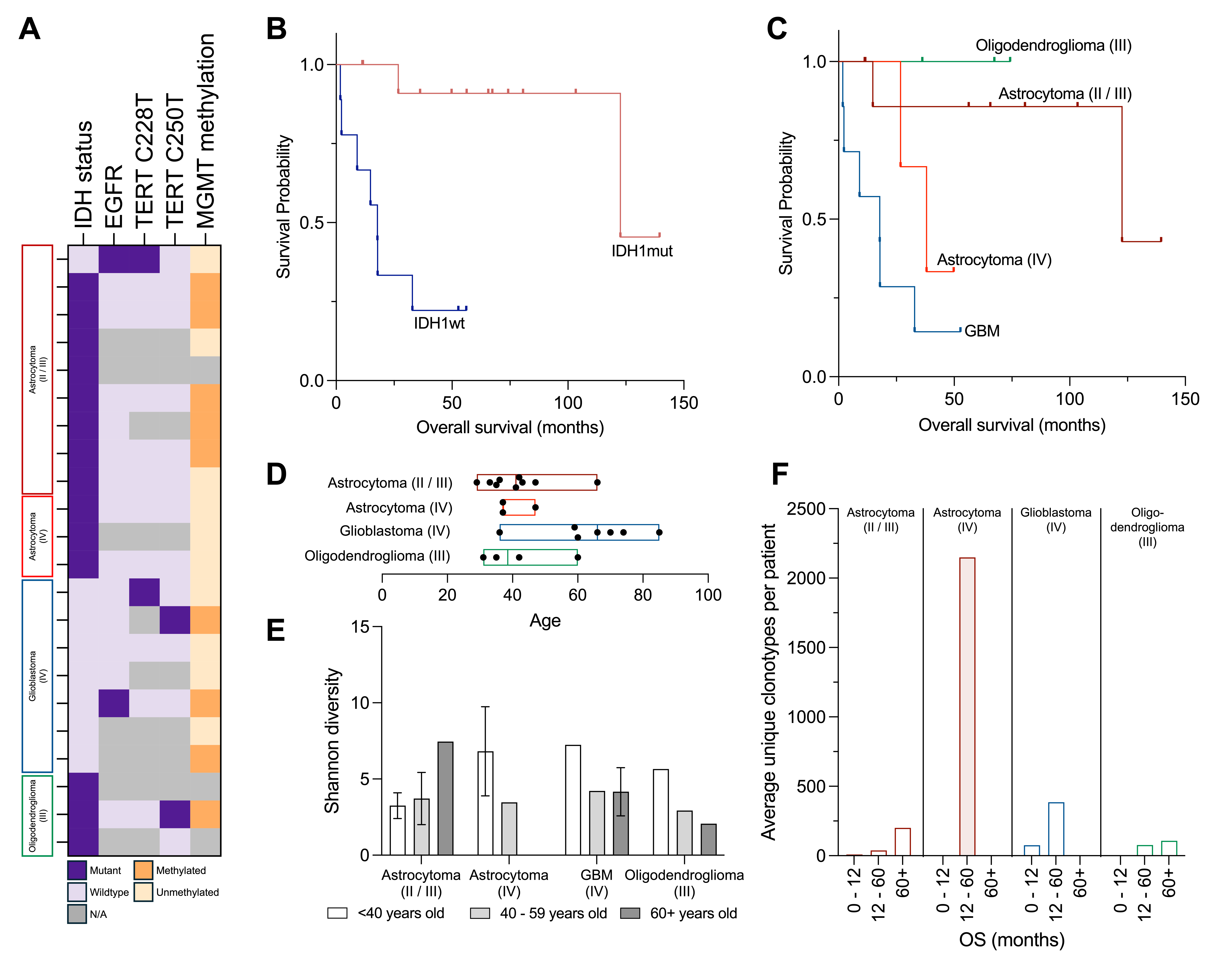

**Supplementary Figure 3. Correlation of TCR diversity and age across patient cohorts.** Tumor mutational burden as determined by histopathology was identified (**A**) to determine potential confounding factors. The overall survival probability of patients with IDH1 mutant (IDH1mut) and IDH1 wildtype (IDH1wt) tumors was determined (**B**) as well as that of each cohort (**C**). Patient age within each cohort (**D**) was defined relative to the average, calculated Shannon Diversity of alpha and beta chains (**E**). The average unique number of clonotypes per patient were calculated for three timeframes of overall survival: 0 – 12 months, 12 – 60 months, and 60+ months (**F**).

**Supplementary Table 1.** Patient sharing in 10 largest, significant clusters across cohorts.

|  | **CD4** | | | | **CD8** | | | |
| --- | --- | --- | --- | --- | --- | --- | --- | --- |
| **Disease** | *# clusters* | *# patients / cluster* | *Average # patient/cluster* | | *# clusters* | *# patients / cluster* | *Average # patient/cluster* | |
| Astrocytoma  (II / III) | 10 | 3 | 3 | (33.3%) | 10 | 3 | 3 | (33.3%) |
| Astrocytoma  (IV) | 10 | 3 | 3 | (100%) | 10 | 3 | 3 | (100%) |
| GBM  (IV) | 10 | 3 | 3 | (42.8%) | 10 | 3 | 3 | (42.8%) |
| Oligodendroglioma  (III) | 10 | 3 | 3 | (100%) | 10 | 3 | 3 | (100%) |
